# Tau Imaging with ^18^F-MK6240 across the Alzheimer’s Disease spectrum

**DOI:** 10.1101/2022.02.13.22270894

**Authors:** Christopher C. Rowe, Vincent Doré, Natasha Krishnadas, Samantha Burnham, Fiona Lamb, Rachel Mulligan, Svetlana Bozinovski, Simon Laws, Regan Tyrell, Kun Huang, Pierrick Bourgeat, Azadeh Feizpour, Olivier Salvado, Colin L. Masters, Jürgen Fripp, Victor L. Villemagne

**Author notes:** Address correspondence to: Professor Christopher C Rowe, Department of Molecular Imaging & Therapy, Austin Health, 145 Studley Road, Heidelberg, Vic. 3084, Australia. **Email:**.

## Abstract

Tau deposition plays a critical role over cognition and neurodegeneration in Alzheimer’s disease (AD). Recent generation tracers have high target to background ratios giving a wide dynamic range that may improve sensitivity for detection of low levels of tau (Pascoal, Shin et al. 2018). Building on previous evidence, this study aims to characterize the effects of tau deposition as assessed by ^18^F-MK6240, in a large cohort of patients across the AD disease spectrum.

A total of 464 participants, enrolled in the AIBL-ADNeT study, underwent ^18^F-MK6240 tau PET, ^18^F-NAV4964 Aβ PET, 3D structural MRI (hippocampal and whole-brain cortical volumes) and extensive neuropsychological evaluation. Participants included 266 cognitively unimpaired controls (CU), 112 patients with mild cognitive impairment (MCI), and 86 patients with probable AD dementia. Evaluation included the characterization of the pattern and degree of ^18^F-MK6240 tracer retention in each clinical group as well as assessment of the relationship between ^18^F-MK6240 and age, Aβ imaging, brain volumetrics and cognition in each of the clinical groups. Standard uptake value ratios (SUVR) were estimated in four predefined composite regions of interest (ROIs), reflecting the stereotypical progression of tau pathology in the brain: 1. Mesial-temporal (Me), 2. Temporoparietal (Te), 3. Remainder of neocortex (R), 4. A temporal meta-region termed metaT+.

^18^F-MK6240 retention was higher in AD patients compared with all other diagnostic groups, with ^18^F-MK6240 distinguishing patients with AD from CU individuals, with the highest effect size obtained in the amygdala (Cohen’s *d*: 2.07), and Me (Cohen’s *d*: 1.99). When considering Aβ status, ^18^F-MK6240 not only was able to distinguish between Aβ+ AD patients and Aβ- CU (Cohen’s *d*: 2.23), but also between Aβ+ and Aβ- CU (Cohen’s *d*: 1.32). In Aβ- CU, ^18^F-MK6240 retention in Me showed a slow age-related increase, while ^18^F-MK6240 retention was higher in younger elderly Aβ+ AD patients compared to their older counterparts. There was a sigmoidal relationship between subthreshold tau and Aβ, providing evidence for a very slow but steady increase in subthreshold tau prior to a fast increase in cortical Aβ. Moreover, a non-linear relationship between Aβ and tau suggest that detectable cortical Aβ precedes detectable cortical tau. While age was the main predictor of cognitive decline in CU, and Aβ and hippocampal volume in MCI, the main predictor of cognitive decline in the AD group was tau. High tau was associated with faster cognitive decline and clinical progression in the CU and MCI groups.

This large study provides further evidence that ^18^F-MK6240 discriminates CU from AD and, most importantly, Aβ+ from Aβ- CU individuals with high effect sizes, suggesting that ^18^F- MK6240 can detect lower tau levels than earlier tau tracers, crucial for early detection of tau deposition as well as tracking small tau changes over time. In conclusion, identification of regional cortical tau deposition has critical diagnostic and prognostic implications and should become a standard tool to identify individuals at risk, as well as outcome measure, in both anti- Aβ and anti-tau trials.

## INTRODUCTION

The term tauopathies designate neurodegenerative conditions, such as Alzheimer’s disease (AD), characterized by the pathological accumulation of tau. In AD, tau hyperphosphorylation leads to tau aggregation in the form of intracellular filamentous inclusions termed neurofibrillary tangles. The mechanisms leading to tau hyperphosphorylation, and aggregation have not yet been fully elucidated, but tau deposition follows a hierarchical stereotypical distribution in the brain (Braak and Braak 1997, Delacourte, David et al. 1999, Serrano-Pozo, Frosch et al. 2011). Aggregated tau is a challenging neuroimaging target, characterized not only by its predominant intracellular location, but also by the different prevalence and combinations of six different tau isoforms. These isoforms, besides being linked to specific phenotypes, are subject to multiple post-translation modifications, leading to heterogeneous ultrastructural conformations of the aggregates (Fitzpatrick, Falcon et al. 2017). In the particular case of AD, these pathological conformations are in much lower concentrations than β-amyloid (Aβ) aggregates in the same brain regions (for in depth review see (Villemagne, Fodero-Tavoletti et al. 2015)).

Since 2011 (Fodero-Tavoletti, Okamura et al. 2011), a steady stream of selective tau tracers for positron emission tomography (PET) have been developed and evaluated in clinical studies (Chien, Bahri et al. 2013, Walji, Hostetler et al. 2016, Stephens, Kroth et al. 2017, Sanabria Bohorquez, Marik et al. 2019, Schmidt, Janssens et al. 2020). The most widely used tracer to date has been ^18^F-flortaucipir (FTP, a.k.a. AV1451 or T807) in the evaluation of AD and non-AD tauopathies (Johnson, Schultz et al. 2016, Scholl, Lockhart et al. 2016), reporting a robust difference in tau tracer retention between cognitively normal elderly controls and AD patients (Cho, Choi et al. 2016, Johnson, Schultz et al. 2016, Wang, Benzinger et al. 2016), as well as in typical and atypical AD presentations where FTP regional retention –not Aβ– matched the clinical phenotype, and also a regional pattern matching Stages 3 and 4 of McKee of Chronic Traumatic Encephalopathy (CTE) (McKee, Stern et al. 2013) in former American Football players (Stern, Adler et al. 2019). In AD, tau tracer retention follows the known distribution of aggregated tau in the brain (Braak and Braak 1997), but also has a close relationship with markers of neuronal injury such as ^18^F-FDG or cortical grey matter atrophy (van Eimeren, Bischof et al. 2017, Xia, Makaretz et al. 2017, Ossenkoppele, Lyoo et al. 2020) while highly predictive of changes in glucose metabolism and grey matter atrophy (Chiotis, Saint-Aubert et al. 2017, La Joie, Visani et al. 2020). Some studies have shown that tau imaging has exquisite differential diagnostic accuracy against other neurodegenerative conditions at the advanced stages of Alzheimer’s disease dementia, (Ossenkoppele, Rabinovici et al. 2018, Leuzy, Smith et al. 2020) and has even been proposed as a “one-stop shop” for the evaluation of dementing neurodegenerative conditions (Hammes, Bischof et al. 2021). To date, all tau tracers have present drawbacks such as “off-target” retention areas in the choroid plexus, basal ganglia, longitudinal sinuses or meninges (Lowe, Curran et al. 2016) and some show strong binding to monoamine oxidase B (MAO-B) instead of tau (Ng, Pascoal et al. 2017). Moreover, some of these tracers present high non-specific binding likely precluding detection of low levels of tau deposition (Baker, Harrison et al. 2019). This might explain why FTP has very high accuracy in detecting Braak stages V-VI (Fleisher, Pontecorvo et al. 2020) while it might be much less accurate at detecting the lower Braak stages.

A recently introduced selective 3R/4R tau tracer, ^18^F-MK6240 (Walji, Hostetler et al. 2016, Pascoal, Shin et al. 2018, Betthauser, Cody et al. 2019), has shown high contrast images with low non-specific binding and low prevalence of off-target binding (Pascoal, Shin et al. 2018, Villemagne, Dore et al. 2018, Betthauser, Cody et al. 2019, Pascoal, Therriault et al. 2020, Kreisl, Lao et al. 2021, Pascoal, Benedet et al. 2021). In a larger cohort of participants, we evaluated 1) the ability of ^18^F-MK6240 to identify lower tau burdens in a large series of cognitively unimpaired (CU) controls, participants with mild cognitive impairment (MCI) and patients with AD dementia, 2) the relationship of ^18^F-MK6240 with age, Aβ, and grey matter atrophy; and 3) the relationship of global and regional ^18^F-MK6240 retention with cognitive performance in the different clinical groups.

## METHODS

### Participants

Four hundred and sixty four participants from the Australian AIBL and ADNeT cohorts were included in the study: 266 cognitively unimpaired controls (CU), 112 participants meeting criteria for mild cognitive impairment (MCI) (Petersen, Smith et al. 1999), and 86 participants meeting NINDS-ADRDA *and* NIAA-AA criteria for AD (McKhann, Drachman et al. 1984, McKhann, Knopman et al. 2011). The AIBL cohort recruitment and evaluation is detailed elsewhere (Ellis, Bush et al. 2009). The study was approved by the Austin Health Human Research Ethics Committee. Written informed consent was obtained from participants and/or from the next of kin or carer for the participants with dementia, prior to participation.

All participants were aged over 50 years, spoke fluent English, and had completed at least 7 years of education. No participants had a history of physical or imaging findings of other neurological or psychiatric illness, current or recent drug or alcohol abuse/dependence, or any significant other disease or unstable medical condition. Between August 2018 and March 2020, participants underwent Aβ PET, tau PET, 3T structural MRI, and extensive neuropsychological examination.

### Neuropsychological evaluation

Participants were administered the Mini Mental State Examination (MMSE), the Clinical Dementia Rating (CDR), Geriatric Depression Scale (GDS), Hospital Anxiety and Depression Scale (HADS), and a battery of neuropsychological tests. The primary cognitive performance measures were the composite memory and non-memory scores, and the Pre-Alzheimer’s cognitive composite for the AIBL cohort (AIBL-PACC) that were calculated as previously described (Pike, Savage et al. 2007, Donohue, Sperling et al. 2014). Briefly, we calculated an Episodic Memory Composite score from the mean of the z-scores (means and standard deviation for creating the z-scores were generated using data from 65 cognitively normal controls with both low ^11^C-PiB PET retention and normal MRI as the reference) for The Rey Complex Figure Test (RCFT, 30 min) Long Delay, the Delayed Recall from the California Verbal Learning Test second edition (CVLT-II), and Logical Memory II. We calculated a Non-Memory Composite score by taking the mean of the z scores for the Boston Naming Test (BNT), Letter Fluency, Category Fluency, Digit Span forwards and backwards, Digit Symbol-Coding, and RCFT Copy (Pike, Savage et al. 2007). The AIBL-PACC score was calculated from the mean of the z-scores of the Delayed Recall from CVLT-II and Logical Memory II, the Digit Symbol substitution test and the MMSE total score (Donohue, Sperling et al. 2014). In participants (n=269, 230 CU, 14 MCI and 25 AD) with available longitudinal neuropsychological data (median [range] 4.8 [1.1-14.4] years), trajectories for the different tests or composites were calculated by linear regression.

### MRI

Participants received an MRI scan using a 3D magnetisation prepared rapid gradient echo (MPRAGE; appendix). The primary MRI performance measures were the cortical grey matter and hippocampal volumes normalised for total intracranial volume. Briefly, MR images for every participant were classified into grey matter, white matter, and CSF using an implementation of the expectation maximisation segmentation algorithm. Hippocampus was extracted using a multi-atlas approach based on the Harmonized Hippocampus Protocol (Boccardi, Bocchetta et al. 2015). Hippocampal volumes were normalised for total intracranial volume, defined as the sum of grey matter, white matter, and CSF volumes. In view of the normal distributions of hippocampal volumes in AIBL healthy controls and Alzheimer’s disease groups, a two-graph receiver operating characteristic curve approach was applied to establish the optimal threshold for hippocampal volume (5.63 cc). Individuals with hippocampal volume of 5.63 cc. or smaller were deemed to exhibit neurodegeneration (N). MRI and psychometric examination were completed within a median [range] of 17 [0-90] weeks of the ^18^F-MK6240 scan.

### PET

#### ^18^F-MK6240 and ^18^F-NAV4698 Synthesis

Labelling was performed in the Department of Molecular Imaging and Therapy, Austin Health, immediately prior to administration. The final product had a radiochemical purity >95%. PET scans were acquired using a Phillips Gemini PET/CT scanner or a Siemens Biograph PET/CT. A CT scan was performed for attenuation correction immediately prior to each imaging period.

#### ^18^F-NAV4694 and ^18^F-MK6240 PET Acquisition

Four 5-min frames were acquired starting at 50 min after injection of 200±15 MBq of ^18^F-NAV4694; and four 5-min frames were acquired starting at 90 min after injection of 184.5±15 MBq of ^18^F-MK6240.

#### Image Analysis

PET scans were analyzed using Computational Analysis of PET from AIBL (CapAIBL^®^) software (Bourgeat, Dore et al. 2018).

For the Aβ imaging studies, the standard Centiloid (CL) method global and a cerebellar regions of interest (ROI) were applied (Klunk, Koeppe et al. 2015). A CL of 25 was used to discriminate high (A+) or low (A-) Aβ burden.

^18^F-MK6240 SUV were used to derive SUVR referenced to the cerebellar cortex at 90-110 min after injection. As previously described (Villemagne, Dore et al. 2017), by grouping the individual ROI, three regional tau composite-masks were constructed: 1. Mesial-temporal (Me) comprising entorhinal cortex, hippocampus, parahippocampus and amygdala; 2. Temporoparietal (Te) comprising inferior and middle temporal, fusiform, supramarginal and angular gyri, posterior cingulate/precuneus, superior and inferior parietal, and lateral occipital; and 3. Remainder of neocortex (R) comprising dorsolateral & ventrolateral prefrontal, orbitofrontal cortex, gyrus rectus, superior temporal, and anterior cingulate. A global measure of neocortical tau was calculated from the average of the ROIs included in the Te and R composites. Additionally, we adopted and adapted a temporal meta-region (TC) described by Jack and colleagues. (Jack, Wiste et al. 2018). While the original composite ROI included entorhinal cortex, amygdala, parahippocampal, fusiform, inferior temporal, and middle temporal regions, we additionally included the hippocampus proper, the temporoccipital region and the gyrus angularis. A 95%ile of the A- CU for each of the respective composite ROIs was used to discriminate high tau from low tau burden. The metaT+ composite was used to define T- or T+ status, because it is a region that allows capture of early neocortical tau deposition beyond the mesial temporal lobe (Villemagne, Lopresti et al. 2020).

### Statistical analysis

Data are presented as mean ± standard deviation (SD) and 95% confidence intervals (CI) unless otherwise stated. Statistical evaluations between groups were performed using a Tukey HSD test. Fisher exact test was used to compare categorical variables. Effect size between CU and AD was measured with Cohen’s *d*. An incremental regression model was used to test if age, sex, years of education, APOE status, Aβ burden, hippocampal volume, and global and regional tau burden significantly predicted changes in cognitive performance. Statistical analysis was performed with JMP for Mac (v13.2, SAS Institute).

### Data availability

Anonymized data may be shared upon request to the corresponding author from a qualified investigator, subject to restrictions according to participant consent and data protection legislation.

## RESULTS

Demographic data are detailed in Table 1. The CU group was older than the AD and MCI groups. As expected, the MCI and AD groups were more cognitively impaired, had a higher prevalence of *APOE e4*, and a higher Aβ burden than CU.

**Table 1.** Demographics.

|  | CU | MCI | AD |
| --- | --- | --- | --- |
| *n* | 266 | 112 | 86 |
| Age | 75.2±5.6 | 72.9±8.1* | 71.2±7.6* |
| Sex (% female) | 57% | 46% | 44% |
| Education (years) | 14.0±3.2 | 11.9±3.4* | 12.0±3.4* |
| *APOE e4* (%) | 31% | 42% | 68%* |
| MMSE | 28.6±1.3 | 26.4±2.3* | 22.8±4.0*† |
| CDR | 0.08±0.2 | 0.48±0.1* | 0.76±0.5*† |
| CDR SoB | 0.12±0.3 | 1.17±0.8* | 4.65±2.3*† |
| Episodic Memory Composite | -0.01±0.7 | -1.72±1.0* | -2.47±10.9*† |
| Non Memory Composite | -0.01±0.6 | -1.03±0.9* | -1.83±1.2*† |
| AIBL-PACC | -0.02±0.7 | -1.83±1.1* | -3.36±1.6*† |
| AIBL-PACC5 v1 | -0.12±0.7 | -1.69±1.0* | -3.08±1.6*† |
| AIBL-PACC5 v2 | -0.01±0.6 | -1.60±1.0* | -3.00±1.5*† |
| Hippocampal Vol (cc) | 5.98±0.5 | 5.58±0.7* | 5.12±0.9*† |
| Cortical Grey Matter Vol (cc) | 463.3±19 | 455.9±23* | 440.5±25*† |
| Aβ burden (CL) | 15.5±34 | 65.8±63* | 93.5±59*† |
| Aβ+ (>25 CL, %) | 17% | 56%* | 80%* |
| Aβ- (CL; n) | 2.3±8 (221) | 3.1±6 (49) | 5.3±7 (17) |
| Aβ+ (CL; n) | 80.2±38 (45) | 114.6±37 (63) | 115.2±44 (69) |

\* Significantly different from CU ( $p < 0.05$ )
† Significantly different from MCI ( $p < 0.05$ )

### Visual inspection

Visual inspection of A- CU cases showed very low ^18^F-MK6240 retention in the brain (Figure 1). In A+ CU cases, tracer retention in MTL was frequently seen, varying in extent from isolated foci in the transrhinal/entorhinal region to a full hook-like distribution involving the entorhinal region, amygdala, hippocampus and parahippocampal regions (“hook sign”). In some cases, there was also involvement of the inferior temporal cortex (Figure 1). In A+ AD cases, high contrast MK retention was more widespread encompassing MTL, lateral occipital, temporoparietal, insula, posterior cingulate/precuneus and prefrontal areas (Figure 1). While A-MCI showed low MK retention in the brain, similar to what was observed in A- CU, A+ MCI cases presented MK retention in MTL and cortical areas involving temporoparietal, temporooccipital, and posterior cingulate. No “off-target” retention in choroid plexus and/or striatum was observed (Figure 1). In about 8-10% of the cases, clear “off-target” retention in meninges was observed, much more noticeable in A- CU and A- MCI cases (Figure 1).

**Figure 1.**
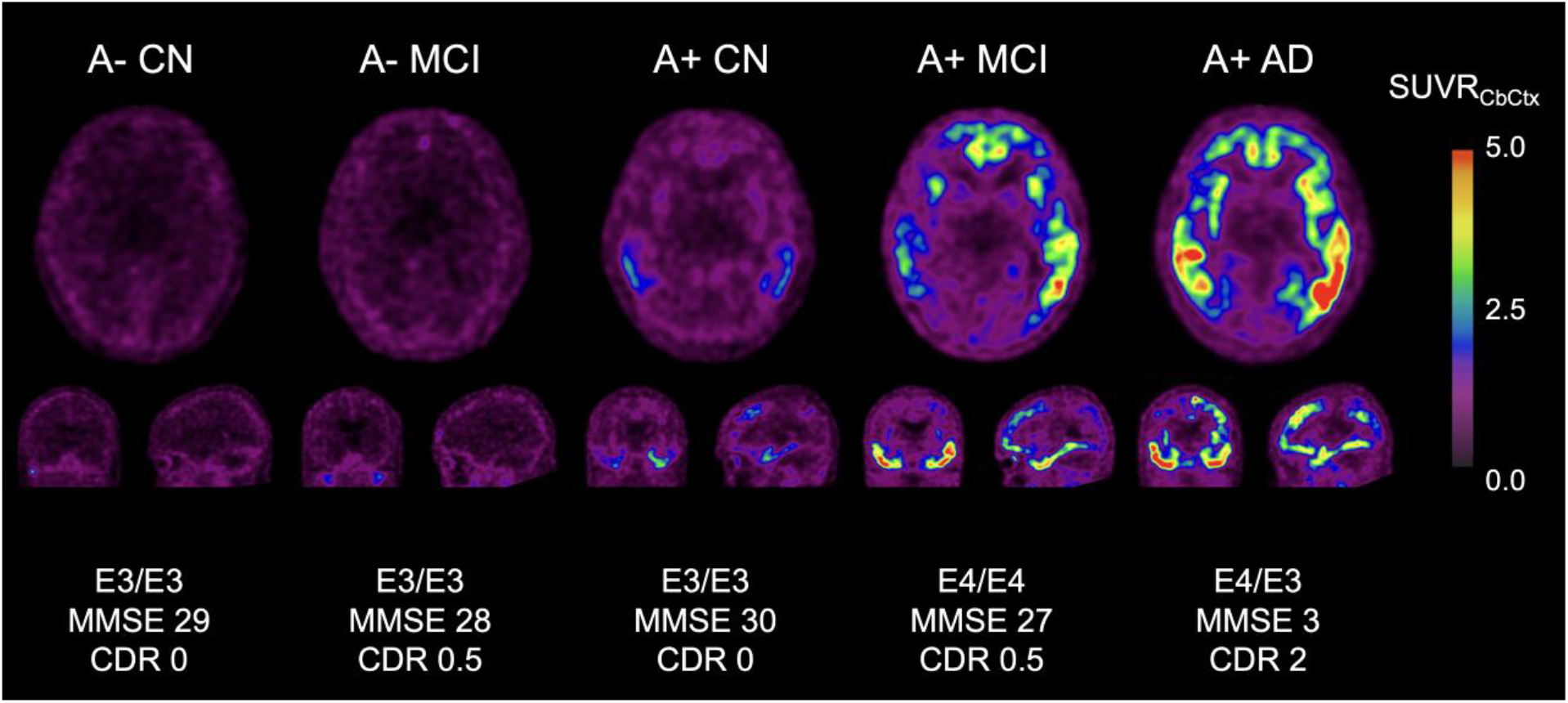
Tau imaging with ^18^F-MK6240 Representative transaxial, coronal and sagittal PET images obtained 90-110 minutes post-injection of participants across the spectrum of clinical diagnosis (aged 65-80) and classified by A status. The A- CU and A- MCI on the left are typical of the appearance of ^18^F-MK6240 images with no cortical binding and very low white matter retention, while A+ MCI and A+ AD are typical high-contrast images showing high tracer retention in neocortical areas. There was no off-target binding in basal ganglia nor choroid plexus, but a “halo” around the brain and cerebellum, due to off-target binding in meninges, was observed.

### Regional and global^18^F-MK6240 retention

Regional and global tau was significantly higher in MCI and AD participants. This difference in ^18^F-MK6240 retention is reflected in very large Cohen’s *d* effect sizes when comparing all CU vs all probable AD, and much larger when comparing all CU to A+ AD (Table 2).

**Table 2.** Regional and global tau by clinical group.

|  | CU | MCI | AD | *Cohen's d* | *Cohen's d* |
| --- | --- | --- | --- | --- | --- |
| *n* | 266 | 112 | 86 | CU vs AD | CU vs A+AD |
| Me | 0.94±0.21 | 1.42±0.60*† | 1.77±0.73* | 1.58 | 1.99 |
| Te | 1.06±0.20 | 1.48±0.67*† | 2.23±1.27* | 1.34 | 1.57 |
| R | 0.90±0.14 | 1.11±0.37*† | 1.58±0.81* | 1.19 | 1.38 |
| metaT+ | 1.03±0.21 | 1.51±0.65*† | 2.07±0.96* | 1.54 | 1.91 |
| Global | 1.00±0.17 | 1.34±0.54*† | 1.98±0.73* | 1.32 | 1.56 |
| Amygdala/Uncus | 0.81±0.31 | 1.55±0.95*† | 2.13±1.14* | 1.61 | 2.07 |
| Hippocampus | 0.84±0.22 | 1.30±0.60*† | 1.62±0.71* | 1.50 | 1.87 |
| Parahippocampus | 0.83±0.17 | 1.13±0.42*† | 1.36±0.49* | 1.50 | 1.81 |
| Entorhinal cortex | 1.28±0.27 | 1.70±0.61*† | 2.01±0.72* | 1.35 | 1.61 |

\* Significantly different from CU ( $p < 0.05$ )
† Significantly different from AD ( $p < 0.05$ )

When separating the clinical groups by Aβ (A) status, and as was observed visually, A+ AD and A+ MCI presented significantly higher SUVR than A- CU in all regions examined (metaT+ shown in Figure 2). ^18^F-MK6240 SUVR was able to discriminate between A- and A+ CU. A+ CU had significantly higher SUVR in metaT+ (Figure 2) and Me, as well as every one of the regions included in the Me composite: hippocampus, amygdala, parahippocampus and entorhinal cortex, resulting in Cohen’s *d* effect sizes ranging between 0.59 in R and 1.32 in the amygdala/uncus (Table 3). As expected, effect sizes were increasingly larger when comparing A- CU to A+ CU (Cohen’s d range 0.59 to 1.32), A- CU to A+ MCI (Cohen’s d range 1.03 to 2.03), and A- CU to A+ AD (Cohen’s d range 1.40 to 2.23) (Table 3). Note that 18% of A+MCI and 13% of A+AD had negative ^18^F-MK6240 tau PET scans (Figure 2).

**Figure 2.**
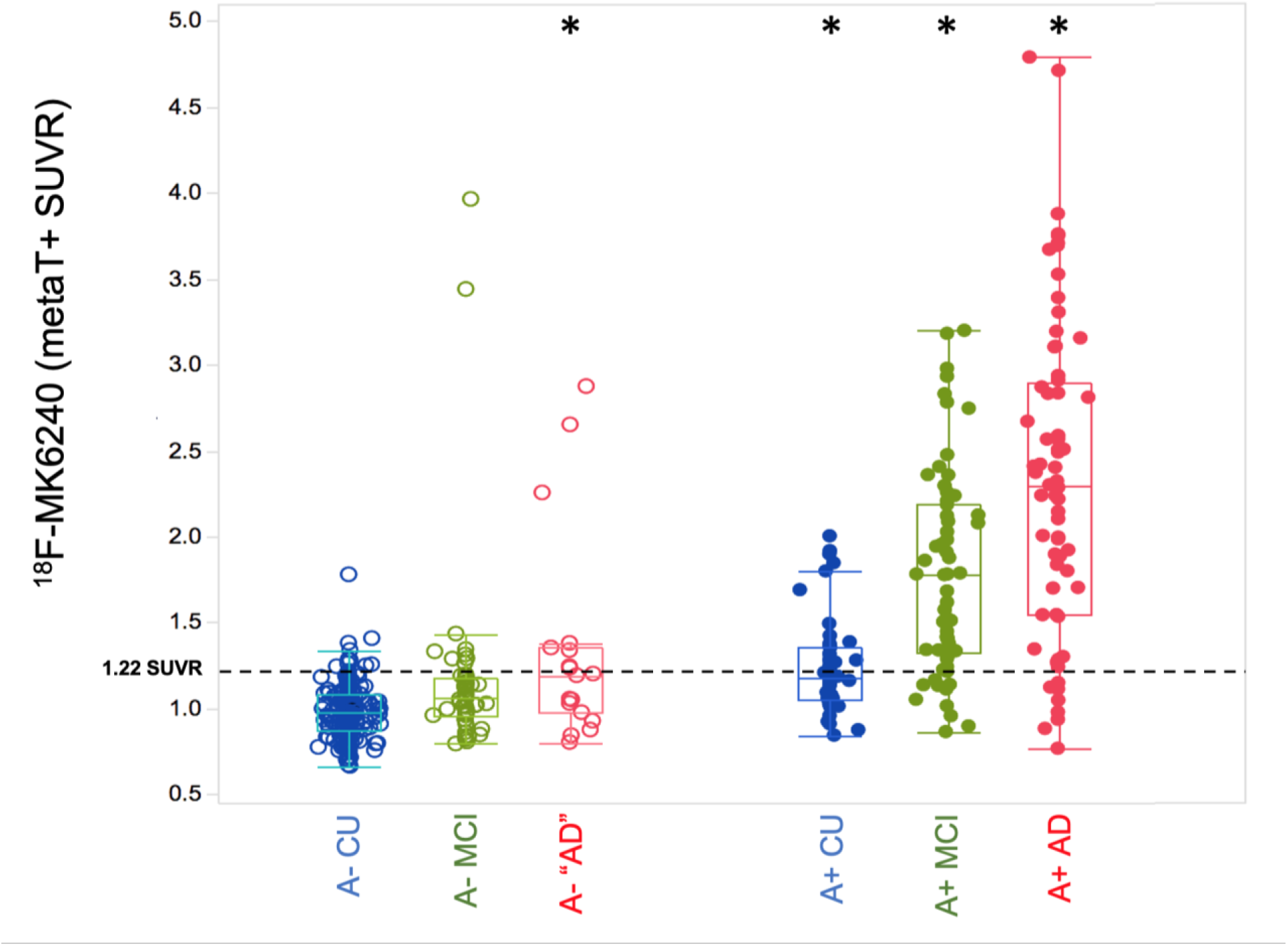
Regional tau by Aβ status as measured by metaT+ ^18^F-MK6240 Box and whiskers plot showing ^18^F-MK6240 metaT+ SUVR across the different clinical groups classified by A status. All A+ groups showed significantly higher metaT+ than A- CU, similar to what was observed on visual inspection, A+ AD and A+ MCI presented significantly higher metaT+ SUVR than A- CU. Furthermore, ^18^F-MK6240 SUVR was able to discriminate between A- CU and A+ CU, where A+ CU presented significantly higher metaT+ SUVR than A- CU. *Significantly different from A- CU (p < 0.05)

**Table 3.** Regional and global tau by clinical group and Aβ status.

|  | A- CU | A+ CU | *d* | A- MCI | A+ MCI | *d* | A+ AD | *d* |
| --- | --- | --- | --- | --- | --- | --- | --- | --- |
| *n* | 221 | 45 | A-CU<br>vs<br>A+CU | 51 | 61 | A-CU<br>vs<br>A+MCI | 68 | A-CU<br>vs<br>A+AD |
| Me | 0.89±0.18 | 1.17±0.26* | 1.24 | 1.06±0.45* | 1.72±0.54* | 2.05 | 1.96±0.70* | 2.11 |
| Te | 1.02±0.13 | 1.22±0.35 | 0.77 | 1.19±0.55 | 1.71±0.68* | 1.41 | 2.47±1.28* | 1.62 |
| R | 0.89±0.12 | 0.98±0.18 | 0.59 | 0.98±0.22 | 1.22±0.44* | 1.03 | 1.72±0.84* | 1.40 |
| metaT+ | 0.98±0.15 | 1.24±0.29* | 1.11 | 1.16±0.54 | 1.79±0.59* | 1.85 | 2.30±0.94* | 1.99 |
| Global | 0.97±0.13 | 1.13±0.28 | 0.74 | 1.11±0.42 | 1.52±0.57* | 1.34 | 2.19±1.08* | 1.60 |
| Amygdala/Uncus | 0.73±0.19 | 1.19±0.46* | 1.32 | 0.93±0.57 | 2.06±1.07* | 2.03 | 2.43±1.07* | 2.23 |
| Hippocampus | 0.80±0.18 | 1.06±0.25* | 1.25 | 0.94±0.43 | 1.60±0.56* | 1.93 | 1.79±0.69* | 1.98 |
| Parahippocampus | 0.80±0.16 | 0.96±0.16* | 0.99 | 0.92±0.35* | 1.31±0.39* | 1.71 | 1.47±0.48* | 1.90 |
| Entorhinal cortex | 1.25±0.25 | 1.45±0.32* | 0.72 | 1.44±0.54 | 1.91±0.58* | 1.49 | 2.15±0.72* | 1.70 |

\* Significantly different from A- CU ( $p < 0.05$ )

When applying the AT(N) classification (Jack, Bennett et al. 2018) to the participants with a clinical diagnosis of probable AD, only 57% were A+T+(N+), 69% were A+T+, 11% were A+T-, 7% were A-T+ (most were in the peri-threshold band for T+), and 13% were A-T-. In CU, 64% were classified as A-T-(N-) (Supplementary Table 1). The MCI group was more evenly split, with 37% being A-T- and 46% A+T+ (Supplementary Table 1).

### Relationship with age

In A- CU, higher age was associated with greater tau in Me (r = 0.22, *p=0*.*0012*) (Figure 3), metaT+ (r = 0.14, *p=0*.*041*), hippocampus (r = 0.24, *p=0*.*0002*), parahippocampus (r = 0.18, *p=0*.*0072*), and amygdala (r = 0.26, *p<0*.*0001*), but no significant associations were observed in neocortical areas. At the prodromal AD stage, younger elderly A+ MCI presented higher tau than their older counterparts only in Me (r = -0.29, *p=0*.*02*) (Figure 3), hippocampus (r = -0.35, *p=0*.*0054*), and amygdala (r = -0.37, *p=0*.*003*), but no significant associations were observed in other areas. This inverse relationship with age was even more evident in A+ AD where younger elderly A+ AD presented higher tau than their older counterparts in Global tau (r = -0.46, *p<0*.*0001*), Te (r = -0.46, *p<0*.*0001*) (Figure 3), R (r = -0.44, *p=0*.*0002*), metaT+ (r = -0.34, *p=0*.*004*), hippocampus (r = -0.28, *p=0*.*019*), and parahippocampus (r = -0.33, *p=0*.*005*), but no significant associations were observed in Me, amygdala or entorhinal cortex. No associations with age were observed in A+ CU, A- MCI, nor A- AD.

**Figure 3.**
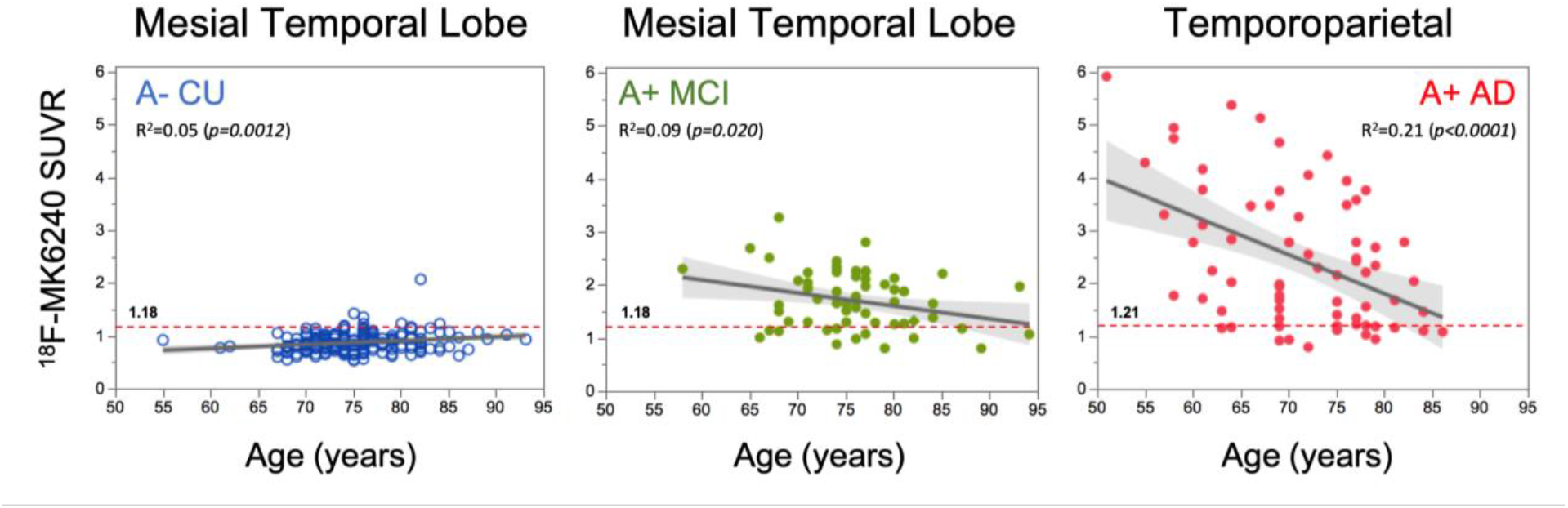
Relationship between mesial temporal and neocortical ^18^F-MK6240 and age in aging and Alzheimer’s disease. In A- CU, there is a slow age-related increase in ^18^F-MK6240 in Me SUVR. Already at the prodromal stage (A+ MCI), this age-related association is inverted. This inverse relationship with age becomes more evident in A+ AD, where younger elderly A+ AD present higher tau than their older counterparts.

### Relationship with Aβ

Cross-sectional analysis of the whole cohort revealed cortical Aβ as measured by ^18^F-NAV4694 CL and tau as measured by ^18^F-MK6240 in metaT+ SUVR were highly, non-linearly correlated (R^2^ = 0.50 (p<0.0001)) (Figure 4). Based on an A and T classification more than half of the participants (56%) were A-T- and 28% A+T+. The rise in Aβ precedes the rise in tau with a slowing in Aβ accumulation, but not in tau, above 100 CL (Figure 4). When tau SUVR are displayed on a logarithmic scale, curve fitting suggests a slow increase in subthreshold tau with increasing Aβ CL with tau crossing the threshold of abnormality once Aβ reaches ∼50 CL (Supplementary Figure 1).

**Figure 4.**
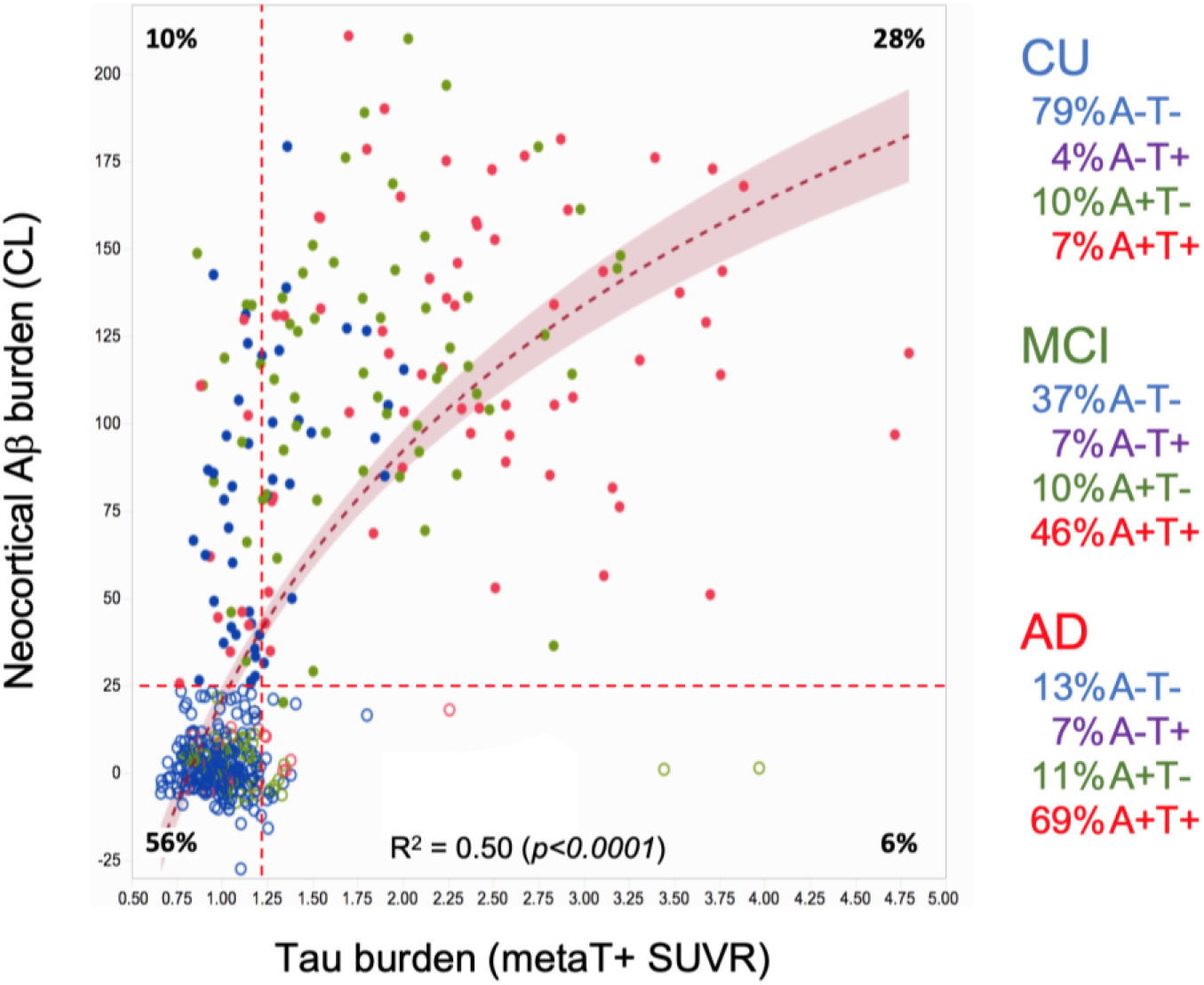
Relationship between neocortical Aβ and tau burden in aging and Alzheimer’s disease. Relationship between neocortical Aβ burden as measured by ^18^F-NAV4694 and expressed in CL, and metaT+ tau burden as measured by ^18^F-MK6240 and expressed in SUVR. A non-linear fit described the association between Aβ and tau (R^2^ = 0.50 (p<0.0001)) better than a linear fit (R^2^ = 0.42 (p<0.0001)). More than half of the participants in this cohort were A-T- and about one third A+T+. Less that 10% of the cases were A-T+. Overall, almost 80% of CU were A-T-, while almost 70% of AD were A+T+. The relationship between Aβ and tau, especially when looking at a region (metaT+) designed to be sensitive to early tau cortical deposition, suggests that detectable increases in Aβ precede detectable increases in tau. Open circles denote A- while full circles denote A+.

After adjusting for age, global and regional tau remained very highly associated with Aβ burden in all regions assessed in the CU and MCI groups, with the highest correlation in metaT+ (R^2^ 0.25; p<0.0001) and lowest in R (R^2^ 0.06; p<0.002). The associations were much lower in AD+, with the highest correlation in the amygdala (R^2^ 0.14; p=0.0017) and lowest in Global tau (R^2^ 0.06; p=0.049).

### Relationship with grey matter volumetrics

After adjusting for age, both global and regional tau showed associations with hippocampal volume (HV) and cortical grey matter volume. In CU, global Aβ burden and tau level in all Me regions and metaT+, were associated with cortical grey matter atrophy. There were no associations between Aβ burden and tau with hippocampal volume. At the MCI stage, global Aβ burden and tau level in all Me regions and metaT+, were also associated with hippocampal atrophy (Figure 5). In AD+, neither global nor regional tau were associated with brain volumetrics, while Aβ only showed a trend (p=0.07) of association with cortical GM. This finding suggests that tau is mainly associated with global atrophy and only in MCI stage we observe a strong association with hippocampal atrophy.

**Figure 5.**
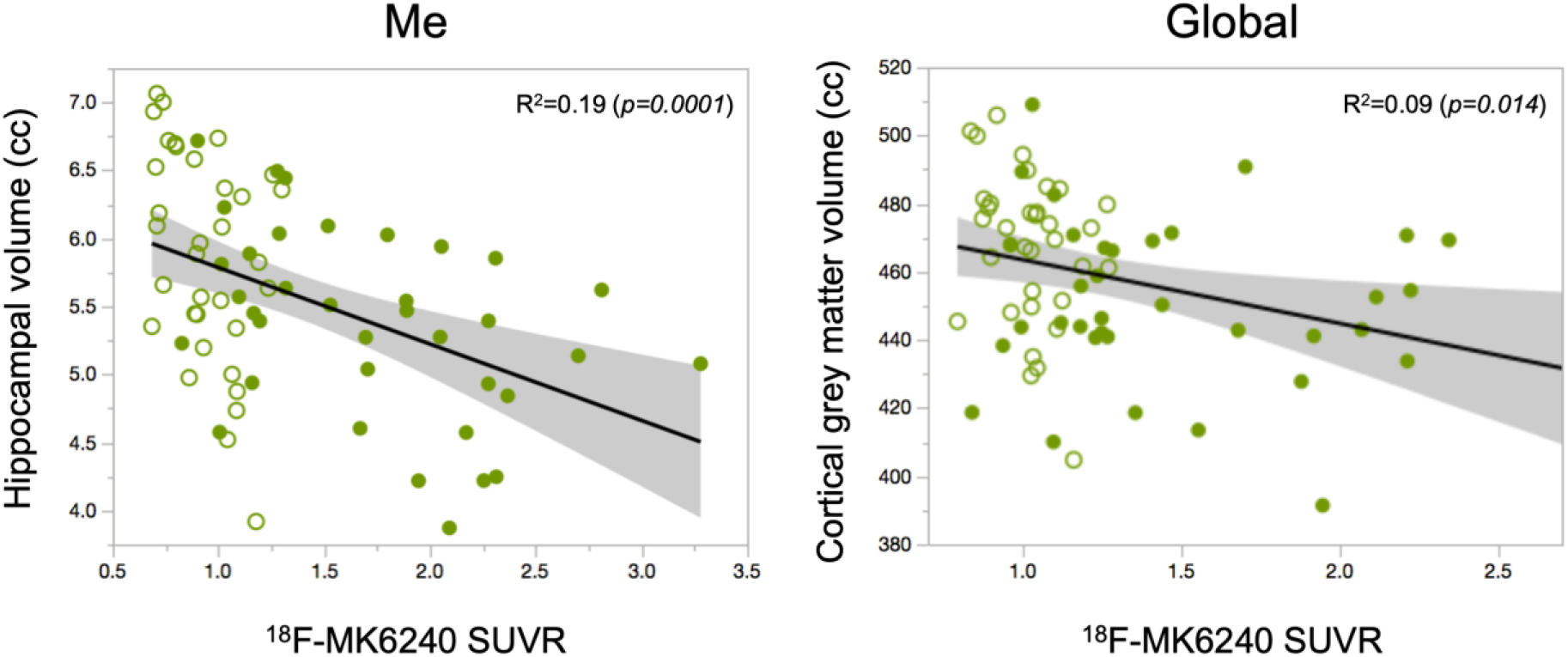
Relationship of mesial temporal ^18^F-MK6240 SUVR with hippocampal volume and global ^18^F-MK6240 SUVR with cortical grey matter volume in MCI. The highest associations between global ^18^F-MK6240 SUVR and cortical grey matter volumes were observed at the MCI stage, which was the only stage showing a relationship between Me ^18^F-MK6240 SUVR and hippocampal volume. Open circles denote A- while full circles denote A+

### Relationship with cognition

When assessing all clinical groups together, metaT+ tau, age, sex, Aβ, and HV were significant predictors (p<0.0001) of performance on some of the cognitive tests (CDR, CDR SoB, Ep Mem). While performance on AIBL-PACC was predicted by sex, Aβ, HV and metaT+ tau, Non-Memory Composite score was predicted only by Aβ, HV and metaT+ tau. Although considering all clinical groups together indicates the *direction* of the association – i.e., the higher the tau levels, the worse the cognitive performance – it does not characterize the independent and/or combined contribution of each of these parameters in relation to cognitive performance in each of the clinical groups.

As expected, MCI and AD participants were significantly more cognitively impaired than CU, irrespective of Aβ status (Table 1). When considering Aβ status, A+ MCI and A+ AD had significantly worse cognitive performance than A- (Supplementary Table 2).

When assessing each clinical group separately, different patterns in the association of tau with cognition were identified. In the CU group, when covariates were taken into account, there were no associations between global and regional tau and cognition, other than Te tau being a weak but significant predictor of change in AIBL-PACC. In this group, the main significant predictors of cognitive performance were age, sex, and years of education, with cognition worsening as a function of increasing age and lower years of education and females performing better than males, especially in tasks requiring memory recall (Table 4). In the MCI group, Aβ, followed by HV and years of education were significantly predictive of changes in cognition. Furthermore, MetaT+ and Me tau had significant associations with Episodic Memory Composite and Te and global tau with Non-Memory Composite (Table 4). A similar pattern was observed in the MCI group when using the AT(N) classification (Supplementary Table 3).

**Table 4.** Relationship between tau and cognition

| **CU** | **MMSE** | **CDR** | **CDR SoB** | **EP MEM** | **NON MEM** | **AIB-PACC** |
| --- | --- | --- | --- | --- | --- | --- |
| | *p* ($\beta$) | *p* ($\beta$) | *p* ($\beta$) | *p* ($\beta$) | *p* ($\beta$) | *p* ($\beta$) |
| Age (yrs) | *n.s.* | *n.s.* | *n.s.* | *n.s.* | 0.0030 (-0.206) | 0.0381 (-0.145) |
| Sex (F) | *n.s.* | 0.0041 (-0.196) | 0.0033 (-0.203) | 0.0003 (0.241) | *n.s.* | <0.0001 (0.262) |
| Years of Education | 0.00401 (0.261) | *n.s.* | *n.s.* | 0.00401 (0.257) | <0.0001 (0.329) | <0.0001 (0.330) |
| APOE4 | *n.s.* | *n.s.* | *n.s.* | *n.s.* | *n.s.* | *n.s.* |
| Aβ burden (CL) | *n.s.* | *n.s.* | *n.s.* | *n.s.* | 0.0257 (0.160) | *n.s.* |
| HV (cc) | *n.s.* | *n.s.* | *n.s.* | *n.s.* | *n.s.* | *n.s.* |
| metaT+ (SUVR) | *n.s.* | *n.s.* | *n.s.* | *n.s.* | *n.s.* | *n.s.* |
| Me (SUVR) | *n.s.* | *n.s.* | *n.s.* | *n.s.* | *n.s.* | *n.s.* |
| Te (SUVR) | *n.s.* | *n.s.* | *n.s.* | *n.s.* | *n.s.* | 0.0389 (-0.138) |
| R (SUVR) | *n.s.* | *n.s.* | *n.s.* | *n.s.* | *n.s.* | *n.s.* |
| Global (SUVR) | *n.s.* | *n.s.* | *n.s.* | *n.s.* | *n.s.* | *n.s.* |
| Amygdala/Uncus (SUVR) | *n.s.* | *n.s.* | *n.s.* | *n.s.* | *n.s.* | *n.s.* |
| Hippocampus (SUVR) | *n.s.* | *n.s.* | *n.s.* | *n.s.* | *n.s.* | *n.s.* |
| Parahippocampus (SUVR) | *n.s.* | *n.s.* | *n.s.* | *n.s.* | *n.s.* | *n.s.* |
| Entorhinal (SUVR) | *n.s.* | *n.s.* | *n.s.* | *n.s.* | *n.s.* | *n.s.* |
| **MCI** | **MMSE** | **CDR** | **CDR SoB** | **EP MEM** | **NON MEM** | **AIB-PACC** |
| | *p* ($\beta$) | *p* ($\beta$) | *p* ($\beta$) | *p* ($\beta$) | *p* ($\beta$) | *p* ($\beta$) |
| Age (yrs) | *n.s.* | *n.s.* | 0.0237 (-0.226) | *n.s.* | *n.s.* | *n.s.* |
| Sex (F) | *n.s.* | *n.s.* | *n.s.* | *n.s.* | *n.s.* | *n.s.* |
| Years of Education | *n.s.* | *n.s.* | 0.0472 (0.192) | 0.0016 (0.276) | *n.s.* | 0.0301 (0.200) |
| APOE4 | *n.s.* | *n.s.* | *n.s.* | *n.s.* | *n.s.* | *n.s.* |
| Aβ burden (CL) | 0.0375 (-0.307) | *n.s.* | <0.0001 (0.609) | 0.0384 (-0.243) | *n.s.* | 0.0036 (-0.371) |
| HV (cc) | *n.s.* | *n.s.* | *n.s.* | 0.0033 (0.286) | *n.s.* | 0.0346 (0.217) |
| metaT+ (SUVR) | *n.s.* | *n.s.* | *n.s.* | 0.0228 (-0.241) | 0.0161 (-0.317) | *n.s.* |
| Me (SUVR) | *n.s.* | *n.s.* | *n.s.* | 0.0143 (-0.290) | *n.s.* | *n.s.* |
| Te (SUVR) | *n.s.* | *n.s.* | *n.s.* | *n.s.* | 0.0015 (-0.377) | *n.s.* |
| R (SUVR) | *n.s.* | *n.s.* | *n.s.* | *n.s.* | *n.s.* | *n.s.* |
| Global (SUVR) | *n.s.* | *n.s.* | *n.s.* | *n.s.* | 0.0044 (-0.338) | *n.s.* |
| Amygdala/Uncus (SUVR) | *n.s.* | *n.s.* | *n.s.* | 0.0147 (-0.322) | *n.s.* | *n.s.* |
| Hippocampus (SUVR) | *n.s.* | *n.s.* | *n.s.* | 0.0100 (-0.304) | *n.s.* | *n.s.* |
| Parahippocampus (SUVR) | *n.s.* | *n.s.* | *n.s.* | 0.0364 (-0.215) | *n.s.* | *n.s.* |
| Entorhinal (SUVR) | *n.s.* | *n.s.* | *n.s.* | *n.s.* | *n.s.* | *n.s.* |

| **AD+** | **MMSE** | **CDR** | **CDR SoB** | **EP MEM** | **NON MEM** | **AIB-PACC** |
| --- | --- | --- | --- | --- | --- | --- |
| | *p* ($\beta$) | *p* ($\beta$) | *p* ($\beta$) | *p* ($\beta$) | *p* ($\beta$) | *p* ($\beta$) |
| Age (yrs) | *n.s.* | *n.s.* | *n.s.* | *n.s.* | *n.s.* | *n.s.* |
| Sex (F) | *n.s.* | *n.s.* | *n.s.* | *n.s.* | *n.s.* | *n.s.* |
| Years of Education | *n.s.* | *n.s.* | *n.s.* | *n.s.* | *n.s.* | *n.s.* |
| APOE4 | *n.s.* | *n.s.* | *n.s.* | *n.s.* | *n.s.* | *n.s.* |
| Aβ burden (CL) | *n.s.* | *n.s.* | *n.s.* | *n.s.* | *n.s.* | *n.s.* |
| HV (cc) | *n.s.* | 0.0033 (-0.444) | 0.0031 (-0.447) | 0.0347 (-0.315) | *n.s.* | *n.s.* |
| metaT+ (SUVR) | 0.0256 (-0.352) | *n.s.* | *n.s.* | *n.s.* | *n.s.* | 0.0063 (-0.429) |
| Me (SUVR) | *n.s.* | *n.s.* | *n.s.* | 0.0279 (-0.332) | *n.s.* | *n.s.* |
| Te (SUVR) | 0.0081 (-0.420) | *n.s.* | *n.s.* | *n.s.* | 0.0024 (-0.483) | 0.0007 (-0.541) |
| R (SUVR) | 0.0042 (-0.450) | *n.s.* | *n.s.* | *n.s.* | 0.0257 (-0.356) | <0.0001 (-0.615) |
| Global (SUVR) | 0.0050 (-0.452) | *n.s.* | *n.s.* | *n.s.* | 0.0038 (-0.464) | 0.0002 (-0.586) |
| Amygdala/Uncus (SUVR) | *n.s.* | *n.s.* | *n.s.* | 0.0434 (-0.306) | *n.s.* | *n.s.* |
| Hippocampus (SUVR) | *n.s.* | *n.s.* | *n.s.* | 0.0228 (-0.328) | *n.s.* | *n.s.* |
| Parahippocampus (SUVR) | *n.s.* | *n.s.* | *n.s.* | 0.0139 (-0.388) | *n.s.* | *n.s.* |
| Entorhinal (SUVR) | 0.0289 (-0.322) | *n.s.* | *n.s.* | *n.s.* | *n.s.* | 0.0281 (-0.327) |

*Items in red denote variables with significant effects ( $p < 0.05$ ) across the different cognitive domains. An incremental regression model including age, sex, years of education, APOE status, Aβ burden, hippocampal volume, and global and regional tau was constructed to assess the independent and/or combined contribution of each parameter over cognitive performance*

AD+ patients presented a completely different pattern, where regional and global tau were the main predictors of cognitive performance (MMSE, Memory and Non-Memory Composite, and AIBL-PACC): while episodic memory was significantly associated with Me tau, MMSE, Non-Memory Composite and AIBL-PACC were significantly associated with cortical tau. In this group, HV was a contributing predictor of change in CDR and CDR SoB (Table 4).

In participants with longitudinal neuropsychological data (median follow up 4.8 years), we looked at the effect of tau over cognitive trajectories and clinical progression. There was a significant faster decline in T+ CU compared to T- CU in all cognitive domains: MMSE (−0.115 vs 0.097, p=0.0009, Cohen’s *d* = -1.0), CDR (0.02 vs 0.004, p=0.0143, Cohen’s *d* = 0.4), CDR SoB (0.069 vs 0.0036, p=0.006, Cohen’s *d* = 0.5), Episodic Memory (−0.071 vs 0.033, p<0.0001, Cohen’s *d* = -0.9), Non-Memory Composite (−0.061 vs -0.004, p=0.0002, Cohen’s *d* = -0.6), and AIBL PACC (−0.082 vs 0.026, p<0.0001, Cohen’s *d* = -0.9). In the MCI group, the same trendwas observed but significance was only reached for MMSE (−0.63 vs 0.016, p=0.035, Cohen’s *d* = -0.8), and CDR SoB (0.70 vs 0.043, p=0.0032, Cohen’s *d* = 4.7).

There were significant differences in clinical progression between T- and T+ in the CU group (p=0.0005), with 18% (6) T+ CU progressing to MCI, 3% (1) to other dementia, and 6% (2) to AD, in contrast to just 4% (9) of T- CU progressing to MCI, and 0.5% (1) to dementia. There were also significant differences between T- and T+ in the MCI group (p=0.0023), with 29% (4) of T- MCI regressing to CU and no one progressing to dementia, whereas 46% (6) of T+ MCI progressed to AD, with no one regressing to CU.

When looking at those with the complete AT(N) classification, 7% of A+T-(N-), 13% of A+T+(N-), 14% of A+T-(N+), and 43% of A+T+(N+) CU participants progressed to MCI, and 11% of A+T+(N+) CU progressed to AD. Only 3% of A-T-(N-), and 3% of A-T-(N+) presented with progression to MCI. Among MCI participants, 33% of A+T+(N-) and 60% of A+T+(N+) progressed to AD.

## DISCUSSION

This study describes the performance of ^18^F-MK6240 to measure tau in the brain, specifically when examined in each clinical group across the AD spectrum. There are several salient features, some related to ^18^F-MK6240 as tracer, and others related to the role of tau as measured by^18^F- MK6240 across the AD spectrum. ^18^F-MK6240 retention in the brain matched the reported histopathological distribution of tau deposits in AD, corresponding to the Braak and Braak or Delacourte stages (Braak and Braak 1997, Delacourte, David et al. 1999, Thal, Rub et al. 2002). ^18^F-MK6240 provides high contrast images with a large dynamic range of SUVR. From a semiquantitative point of view, it has been previously shown that SUVR has a high correlation with BP_ND_ and distribution volume ratio (DVR) (Pascoal, Shin et al. 2018, Betthauser, Cody et al. 2019) and a test-retest variability <7% (Salinas, Lohith et al. 2018). While off-target retention in meninges was observed in 8-10% of the cases, there was no off-target binding in choroid plexus and striatum as observed with FTP and, to a lesser degree, with ^18^F-GTP-1 (Sanabria Bohorquez, Marik et al. 2019) and ^18^F-RO948 (Leuzy, Smith et al. 2020). The absence of off-target binding in the choroid plexus is important because it allows the examination of MTL structures like the hippocampus without having to account for the potential spill-over from the choroid plexus or, in order to avoid performing complex corrections, discard this region that plays a critical role in AD from the analysis. As with most tau imaging tracers, there was also off-target retention in anterior midbrain, likely reflecting specific (saturable) binding to melanin (Marquie, Normandin et al. 2015).

The protracted but steady age-related increase in MTL SUVR in A- CU, along with younger elderly A+ AD presenting with higher tau levels compared to their older A+ AD counterparts, have been reported before with other tracers (Pontecorvo, Devous et al. 2017) as well as ^18^F- MK6240 in smaller samples (Betthauser, Koscik et al. 2020, Kreisl, Lao et al. 2021). Our study shows that this age-related observation at the AD stage is already starting to manifest in prodromal AD (A+ MCI) cases.

The relationship between tau and Aβ has been also repeatedly reported with other tau and Aβ tracers (Johnson, Schultz et al. 2016, Pontecorvo, Devous et al. 2017) and more recently with ^18^F-MK6240 (Betthauser, Koscik et al. 2020, Pascoal, Therriault et al. 2020). We found that a non-linear fit best explains the data, showing that a rise in detectable Aβ precedes a detectable increase in tau. We have previously shown that Aβ levels above ∼50 CL are required to show significant cognitive and clinical change in cognitively normal older persons followed for 8 years (van der Kall, Truong et al. 2020). ^18^F-MK6240 tau PET has shown that this ∼50 CL level is the point where a significant rise in the prevalence of T+ cases in CU as well as MCI and AD patients occurs (Dore, Krishnadas et al. 2021).

The temporal meta-region (Jack, Wiste et al. 2018) provides a sensitive way to detect early tau deposition and to yield the highest discriminatory accuracy between AD and non-AD neurodegenerative conditions (Ossenkoppele, Rabinovici et al. 2018). Given that off-target binding in choroid plexus is not observed with ^18^F-MK6240, our modified version of Jack’s temporal meta-region includes the hippocampus proper, while also adding the temporooccipital, angular and supramarginal gyri, areas of early cortical Aβ and tau deposition. As we previously reported (Villain, Chetelat et al. 2012), and was recently confirmed (Guo, Landau et al. 2020), the supramarginal gyrus is an area of early Aβ deposition. Consequently the metaT+ ROI is likely well suited to document the transition from primary age related tauopathy (PART) to AD, and will capture the regional tau patterns described for the different tau pathological subtypes in AD including limbic predominant and hippocampal sparing variants (Murray, Graff-Radford et al. 2011, Charil, Shcherbinin et al. 2019). For these reasons, we selected the metaT+ region to define T+ in this analysis (Villemagne, Lopresti et al. 2020).

^18^F-MK6240 yielded high Cohen’s d effect sizes between CU, MCI, and AD, that were even higher when classifying by A status. Much more interesting was the ability of ^18^F-MK6240 to discriminate between A- and A+ CU in Me and metaT+, suggesting ^18^F-MK6240 is able to discriminate individuals with low levels of tau deposition, with Cohen’s *d* effect sizes ranging between 0.59 in R and 1.32 in the amygdala/uncus. High sensitivity to detect low tau levels is crucial when using tau imaging for participant selection, monitoring and outcome measure in anti-Aβ trials (Mintun, Lo et al. 2021). When applying the AT(N) framework (Jack, Bennett et al. 2018) (Supplementary Table 1), a clear categorical separation between those participants with AD or AD pathophysiology and those without AD became evident.

While the overall trend shows that higher cortical tau is associated with lower cognitive performance, when assessed in detail by clinical group, tau, as well as Aβ, seem to be necessary but not sufficient to explain all the cognitive impairment variance. Factors of resilience and susceptibility also play an important role because A+ CU presented higher tau than A-CU while showing no cognitive impairment. A previous study, using mediation analysis, showed that Aβ accumulation leads to tau accumulation and this in turn leads to cognitive impairment in CU (Hanseeuw, Betensky et al. 2019). Moreover, we believe that the assessment of synaptic and neuronal injury, the (N) in the new classification framework (Jack, Bennett et al. 2018) is the missing extra variable between tau and cognitive impairment. We can see this at the MCI stage, before all parameters reach a floor. If we use the AT(N) classification to compare A-T+(N-) MCI to A-T+(N+) MCI, those (N+) have significantly lower AIBL PACC performance than (N-) (− 0.72±0.7 vs -2.81±1.0, respectively, p=0.04) (Supplementary Table 3). And it is at the MCI stage that we observe a tight relationship between tau burden in Me and hippocampal volume, while global tau correlates with cortical grey matter atrophy.

When assessing the different factors associated with cognitive performance in different domains, we identify different patterns in each clinical group. In AD+ patients, tau was the main predictor of cognitive impairment. Episodic Memory was significantly associated with MTL tau. MMSE, Non-Memory Composite, and AIBL-PACC were significantly associated with cortical tau. CDR and CDR SoB were associated with HV. In the MCI group, the main associations with cognitive performance were Aβ, HV and years of education. In MCI we also observed the effect of tau location on specific cognitive domains. In MCI metaT+ and MTL tau were significantly associated with Episodic Memory Composite. Te and global tau were significantly associated with Non-Memory Composites. In CU, significant associations with cognitive performance were age, sex, and years of education with worse cognitive performance associated with increasing age, lower years of education and females performing better than males, especially in tasks requiring memory recall. Also in CU, Aβ burden only was associated with the Non-Memory Composite. Overall, the strongest effect of tau is observed at the AD stage, likely because tau is higher, and other potential drivers of cognitive decline, like Aβ, have already reached a plateau. Longitudinal cognitive assessment showed high tau was associated with faster cognitive decline and with clinical progression to MCI or AD in the CU and MCI groups confirming previous reports (Betthauser, Koscik et al. 2020, Ossenkoppele, Smith et al. 2021). Faster progression is observed in A+T+ participants compared to A+T-.

There are distinct limitations of this study. While SUVR is highly associated with DVR, at higher levels of tau, ^18^F-MK6240, like most of the other tau tracers, does not reach apparent steady state and may under-estimate the total tau burden. The off-target tracer retention in meninges could affect ^18^F-MK6240 quantification though because the degree of meningeal ^18^F-MK6240 retention is similar around cortical regions as in the cerebellar cortex reference region, the potential influence of spill-over may cancel out. We compared results before and after applying a meningeal mask to avoid sampling meninges, and we further applied partial volume correction to account for the potential meningeal spill-over, and there were no significant differences in the SUVR results (data not shown). Because the issue of the effect of meningeal retention on semiquantitative measures is beyond the scope of the present study, meningeal results will be presented in a separate paper. While we accounted for several variables (age, sex, Aβ, hippocampal atrophy) when assessing the effects of tau on cognitive performance, there were others that were not considered, such as APOE status or cardiovascular risk that might contribute independently to cognitive impairment. APOE status was only available in 67% of the cohort and its introduction in the model reduced the sample size assessed without adding information beyond what was provided by Aβ. Moreover, in view of the stringent selection criteria for the study, individuals with cerebrovascular disease or other comorbidities were excluded, so their effect on cognitive performance is likely underestimated. Lastly, the participants were volunteers who were not randomly selected from the community, were generally well-educated and had high scores on cognitive tests. Thus, these findings regarding cognitive performance might only be valid in similar cohorts, precluding the generalisation of the findings to the general population.

## CONCLUSION

^18^F-MK6240 provides images of high target to background contrast without “off-target” binding in basal ganglia or choroid plexus, although mild midbrain and meningeal tracer retention was observed in ∼10% of cases. The cortical binding of ^18^F-MK6240 provided discrimination of individuals with clinical diagnosis of AD from cognitively unimpaired elderly participants, but most importantly between A- CU and A+ CU suggesting ^18^F-MK6240, in contrast to reports with other tau tracers, can robustly detect low levels of tau deposition at the preclinical stages of AD. This is important from a staging and prognosis and has implications for the identification of individuals at risk for disease specific therapeutic trials and assessing longitudinal tau changes over time.

The separate assessment of the relationship between tau and cognitive performance in the different clinical groups, allowed a better dissection of the role of tau and cognition along the AD spectrum, showing that cortical tau is the main predictor of cognition at the AD stage. The implementation of ^18^F-MK6240 studies promises to be a significant step towards the integration of tau imaging into clinical practice and towards meeting the desired goal of earlier diagnosis of AD to assist the development of preventative interventions and therapeutic trials.

## ACKNOWLEDGEMENTS

The authors would like to thank the Brain Research Institute for support in acquiring the MRI data. Some of the data used in the preparation of this article was obtained from the Australian Imaging Biomarkers and Lifestyle flagship study of aging (AIBL), and the Australian Dementia Network (ADNeT) that receive funding support from the National Health and Medical Research Council (NHMRC). We also thank the participants who took part in the study and their families.

