## Supplementary Table 1; Supplementary Table 2; Supplementary Table 3; Supplementary Figure 1 for "Tau Imaging with ^18^F-MK6240 across the Alzheimer’s Disease spectrum"

**SUPPLEMENTARY MATERIALS**

**Supplementary Table 1. AT(N) Classification**

|  | **CN** | **MCI** | **AD** |
| --- | --- | --- | --- |
| *n* | *231* | *100* | *76* |
| **A-T-N-** | 64% | 21% | 4% |
| **A-T-N+** | 17% | 16% | 9% |
| **A-T+N-** | 3% | 4% | 4% |
| **A-T+N+** | 1% | 4% | 3% |
| **A+T-N-** | 7% | 5% | 5% |
| **A+T-N+** | 3% | 6% | 5% |
| **A+T+N-** | 4% | 15% | 13% |
| **A+T+N+** | 1% | 31% | 57% |

**Supplementary Table 2. Cognitive performance by Aβ status**

|  | **A- CU** | **A+ CU** | **A- MCI** | **A+ MCI** | **A+ AD** |
| --- | --- | --- | --- | --- | --- |
| *n* | *221* | *45* | *51* | *61* | *68* |
| MMSE | 28.7±1.2 | 28.2±1.4 | 27.1±2.3*† | 25.8±2.3*†# | 22.6±4.4* |
| CDR | 0.07±0.2 | 0.11±0.2 | 0.48±0.1*† | 0.49±0.1*† | 0.80±0.5* |
| CDR SoB | 0.12±0.4 | 0.14±0.3 | 0.94±0.7*† | 1.38±0.8*† | 4.84±2.5* |
| Ep Memory | -0.00±0.7 | -0.07±0.7 | -1.31±0.9*† | -2.03±1.0*†# | -2.44±0.9* |
| Non-Memory | 0.01±0.6 | -0.06±0.7 | -0.85±0.9*† | -1.19±0.8*† | -1.82±1.2* |
| AIBL-PACC | -0.00±0.7 | -0.13±0.8 | -1.35±1.0*† | -2.21±1.0*†# | -3.43±1.7* |

** significantly different from CU- (p<0.05)*

*† significantly different from AD+ (p<0.05)*

*# significantly different from MCI- (p<0.05)*

**Supplementary Table 3. Cognitive performance by AT(N) status**

| **CN** | A-T-N- | A-T-N+ | A-T+N- | A-T+N+ | A+T-N- | A+T-N+ | A+T+N- | A+T+N+ |
| --- | --- | --- | --- | --- | --- | --- | --- | --- |
| *n* | *147* | *39* | *7* | *3* | *16* | *6* | *10* | *3* |
| MMSE | 28.8±1.1 | 28.4±1.5 | 28.9±0.9 | 27.7±1.2 | 28.5±1.2 | 28.5±1.6 | 27.9±1.9 | 28.7±0.6 |
| CDR | 0.06±0.2 | 0.06±0.2 | 0.00±0.0 | 0.33±0.3 | 0.06±0.2 | 0.08±0.2 | 0.15±0.2 | 0.00±0.0 |
| CDR SoB | 0.12±0.4 | 0.13±0.3 | 0.00±0.0 | 0.33±0.3 | 0.06±0.2 | 0.08±0.2 | 0.25±0.4 | 0.00±0.0 |
| EP MEM | 0.05±0.7 | 0.05±0.7 | -0.03±0.9 | -0.58±0.3 | 0.14±0.6 | 0.22±0.9 | -0.15±0.7 | 0.04±0.3 |
| NON-MEM | 0.06±0.6 | -0.09±0.6 | -0.14±0.3 | -0.68±0.3 | 0.19±0.7 | 0.01±0.7 | -0.03±0.6 | 0.06±0.3 |
| AIBL-PACC | 0.06±0.7 | -0.07±0.7 | -0.12±0.6 | -0.45±0.6 | 0.17±0.6 | 0.07±1.1 | -0.34±0.7 | 0.04±0.3 |
| **MCI** | A-T-N- | A-T-N+ | A-T+N- | A-T+N+ | A+T-N- | A+T-N+ | A+T+N- | A+T+N+ |
| *n* | *21* | *16* | *4* | *4* | *4* | *5* | *15* | *31* |
| MMSE | 27.3±1.8 | 27.6±2 | 28.8±1.3 | 23.8±2.9 | 27.8±1.7 | 26±2.4 | 25.5±2.1 | 25.4±2.4 |
| CDR | 0.45±0.2 | 0.50±0.0 | 0.50±0.0 | 0.50±0.0 | 0.50±0.0 | 0.50±0.0 | 0.50±0.0 | 0.50±0.0 |
| CDR SoB | 0.86±0.5 | 0.72±0.4 | 1.13±0.8 | 0.63±0.6 | 1.25±0.9 | 1.3±0.6 | 1.13±0.7 | **1.50±0.8*** |
| EP MEM | -1.20±1.0 | -1.17±0.8 | -1.30±1.2 | -2.04±1.2 | -1.33±0.6 | -2.13±1.0 | -1.62±0.9 | **-2.46±0.9*** |
| NON-MEM | -0.93±0.8 | -0.53±0.7 | -0.23±0.5 | -1.77±1.9 | -1.03±1.3 | -1.06±0.4 | -1.16±0.8 | -1.22±0.9 |
| AIBL-PACC | -1.16±0.8 | -1.21±0.9 | -0.71±0.7 | -2.81±1.0 | -1.34±0.9 | -2.16±1.0 | -1.87±1.1 | -**2.58±1.0*** |
| **AD** | A-T-N- | A-T-N+ | A-T+N- | A-T+N+ | A+T-N- | A+T-N+ | A+T+N- | A+T+N+ |
| *n* | *3* | *7* | *3* | *2* | *4* | *4* | *10* | *43* |
| MMSE | 23.7±3.1 | 23.7±2.4 | 23.0±1.0 | 21.5±0.7 | 25.5±1.7 | 24.8±3.6 | 23.9±2.3 | 22.0±5.0 |
| CDR | 0.67±0.3 | 0.71±0.3 | 0.67±0.3 | 0.50±0.0 | 0.63±0.3 | 0.88±0.8 | 0.60±0.2 | 0.83±0.5 |
| CDR SoB | 4.00±0.9 | 3.71±1.3 | 4.00±0.5 | 4.50±0.7 | 3.63±0.9 | 5.38±3.1 | 3.95±1.0 | 5.05±2.7 |
| EP MEM | -1.81±0.7 | -2.91±0.6 | -2.31±0.6 | -3.47±0.0 | -1.29±1.2 | -2.15±0.8 | -2.44±0.5 | -2.51±0.9 |
| NON-MEM | -2.61±2.1 | -1.66±1.5 | -2.46±0.7 | -2.24±1.3 | -1.67±1.2 | -1.05±0.3 | -1.80±1.2 | -1.89±1.2 |
| AIBL-PACC | -2.86±1.6 | -2.83±0.7 | -3.06±0.3 | -3.62±0.1 | -1.76±1.0 | -2.39±1.4 | -2.85±0.8 | -3.78±1.9 |

** significantly different from MCI- (p<0.05)*

**Supplementary Figure 1**. Relationship between neocortical Aβ and tau burdens in aging and Alzheimer’s disease.

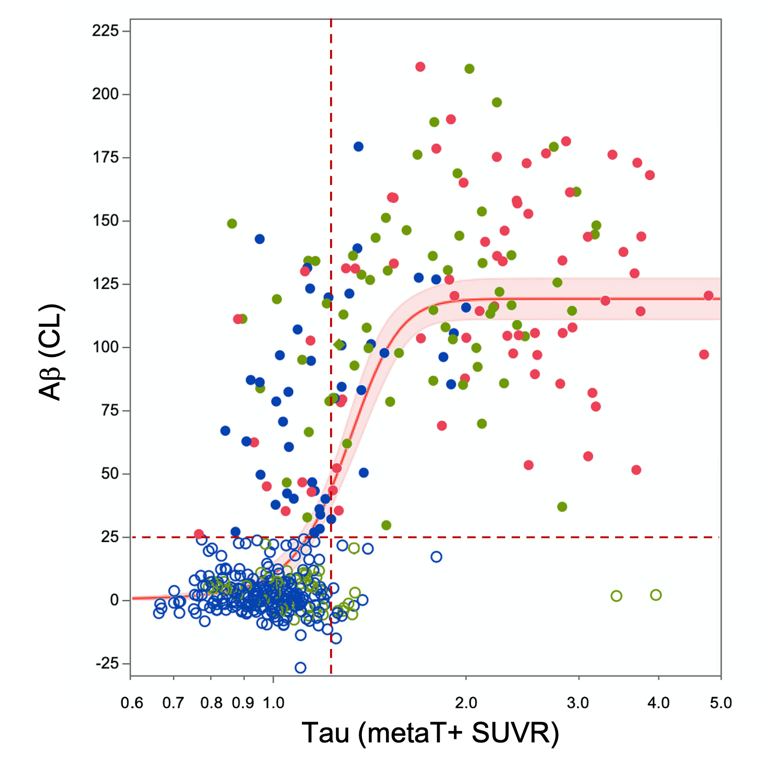

*When tau SUVR are displayed using a logarithmic scale, curve fitting reveals a sigmoidal relationship between tau and Aβ, where a very slow but steady increase in subthreshold tau precedes a fast increase in cortical Aβ that is then followed, as Aβ tends to a plateau, by a steady increase in cortical tau. Open circles denote A- while full circles denote A+*
